# Effects of an exercise prescription system (ACTIVE) on physical function in community-dwelling older adults: protocol for a single-centre, assessor-blinded, parallel-group randomised controlled trial

**DOI:** 10.64898/2026.11.20.26352522

**Authors:** Yao Pan, Hongxin Liu, Yan Li, Yuang Hao, Meng Qing, Zimin Wang, Xin Miao, Yan Xu

## Abstract

**Background:** As China’s population continues to age, preserving physical function in older adults is an important goal of healthy ageing. Digital health interventions have been used to promote physical activity. Comparative evaluation of specific programmes is needed to determine whether individualised exercise prescriptions informed by multidimensional functional assessment provide additional functional benefits.

**Methods:** This single-centre, assessor-blinded, parallel-group randomised controlled trial planned sample size of 142 community-dwelling adults aged 60–85 years in Beijing. Participants will be allocated 1:1 to the ACTIVE group or the standardised, non-individualised exercise group (71 per group) for an 8-week intervention. The primary outcome is the physical function composite Z-score at week 8. It is the unweighted mean of four measures after aligning their directions and standardising them against baseline values: 4-m gait speed, five-times sit-to-stand time, 6-min walk distance and eyes-open single-leg stance time. Secondary outcomes include the raw values and Z-scores of the four components, physical activity measured with the International Physical Activity Questionnaire Short Form (IPAQ-SF), health-related quality of life measured with the EQ-5D-5L, chair sit-and-reach performance, lower-limb muscle strength and the strength asymmetry index, and handgrip strength.

**Discussion:** This trial will evaluate the short-term effects of ACTIVE on physical function in community-dwelling older adults, providing preliminary clinical evidence for assessment-informed individualised home-based exercise and informing subsequent research and community implementation.

**Trial registration:** Chinese Clinical Trial Registry (ChiCTR2600117481). Registered on 26 January 2026. https://www.chictr.org.cn/

## Introduction

According to China’s Seventh National Population Census, adults aged 60 years and older accounted for 18.70% of the population in 2020.^1^ Maintaining the physical function needed to live independently is a goal of healthy ageing.^2^ Poorer lower-extremity performance in community-dwelling older adults is associated with a higher risk of subsequent disability.^3^ Even among generally healthy older adults, distinct patterns of impairment in intrinsic capacity are associated with possible sarcopenia and fall risk.^4^ Identifying changes in physical function and intervening before marked limitations develop offer an important opportunity to delay functional decline and disability.^5^ Exercise is widely regarded as one of the most cost-effective strategies for maintaining physical function.^6^ Its benefits depend on participation and on how well the exercise content, starting dose and progression match the individual’s functional capacity.^7,8^

Randomised controlled trials support the use of multicomponent exercise to improve or maintain physical function in older adults. The REACT trial showed that exercise combined with behavioural support helped maintain lower-limb function in older adults at risk of mobility limitation.^9^ In the LIFE trial, structured physical activity reduced the risk of major mobility disability compared with health education.^10^ The Spanish Vivifrail trial assigned home-based multicomponent exercise according to functional level in frail or prefrail older adults with mild cognitive impairment or mild dementia. Short Physical Performance Battery (SPPB) scores improved compared with usual care.^11^ Chen and colleagues also observed improved SPPB scores following multicomponent exercise in older adults with functional decline.^12^ These studies primarily enrolled older adults with mobility limitations, frailty or functional decline, or those living in institutional settings. The comparators were generally usual care or health education. The evidence therefore supports multicomponent exercise in older adults at increased risk of functional decline. Whether it improves overall physical function in community-dwelling older adults with relatively preserved function and whether individualised prescription adds benefit over standardised exercise remain open questions.

Given the established benefits of exercise,^13,14^ appropriate exercise prescription is central to further improving physical function. International recommendations for older adults advise selecting exercise type, frequency, duration and intensity according to functional capacity and exercise tolerance, with gradual adjustments based on functional performance.^7,8^ Vivifrail applied this principle by assigning training levels based on SPPB performance.^11^ StandingTall used balance ability and self-reported exertion to guide progression.^15^ These studies illustrate the use of baseline assessment and feedback in exercise prescription, although the range of domains assessed was relatively limited. For programmes targeting muscle strength, flexibility, balance, coordination and cardiorespiratory endurance, it remains unclear how best to translate multidimensional assessments into exercise modules, starting doses and rules for progression and regression, and whether this degree of individualisation offers additional functional benefits.

Digital technology can support delivery of this approach at home. Li and colleagues found that a mobile health programme delivering lifestyle-integrated multicomponent exercise improved frailty status in prefrail community-dwelling older adults compared with health education.^16^ The Australian StandingTall trial delivered individualised balance training through a tablet application to 503 independently living older adults and adjusted the difficulty according to self-reported exertion. Standing balance improved at 6 and 12 months compared with health education. At 12 months, the groups did not differ significantly in the fall rate or the proportion of participants who fell.^15^ These studies show that digital platforms can deliver home-based exercise, monitor participation and collect feedback. Health education has commonly served as the comparator. The added benefit of individualised prescription when both groups use the same digital platform and a common standardised exercise framework remains uncertain.

REACT, LIFE, Vivifrail and the Chinese multicomponent exercise trial primarily enrolled older adults with mobility limitations, functional decline or a risk of frailty.^9–12^ StandingTall enrolled independently living community-dwelling older adults and focused on balance and fall prevention.^15^ The present trial focuses on healthy older adults. Proactive health and primary prevention emphasise identifying and addressing health risks early to maintain physical function before disability develops.^17,18^ Offering exercise to healthy older adults before substantial functional impairment develops may help delay functional decline, preserve independent living and reduce future care needs at the population level.

We developed ACTIVE, a decision system that uses knowledge-based rules to prescribe exercise. It integrates baseline assessments of lower-limb strength, sarcopenia status, cardiorespiratory endurance, muscle strength asymmetry and functional movement performance. Prespecified thresholds convert assessment results into structured labels that guide the selection of exercise modules, the starting dose and the level of difficulty. During training, the system uses participant-reported difficulty to progress, maintain or regress the exercises. The rules were specified before the trial and remain fixed throughout it, with no online learning or updates. ACTIVE translates the assessment and progression principles in exercise recommendations,^7,8^ together with the use of functional stratification and perceived exertion to adjust difficulty in earlier programmes,^11,15^ into explicit rules for module selection, starting dose and difficulty adjustment. This approach reduces repetitive manual decisions while retaining professional guidance and monitoring. Using the same digital platform (a WeChat mini programme) and the same basic support in both groups, this trial will compare ACTIVE-guided individualised multicomponent exercise with standardised, non-individualised multicomponent exercise in community-dwelling adults aged 60–85 years. The comparison will determine whether baseline assessment and feedback-based adjustments yield additional benefit on the physical function composite Z-score at week 8.

### Objective and hypothesis

This study aims to compare the effects of an 8-week ACTIVE programme and a standardised, non-individualised exercise programme on physical function in community-dwelling adults aged 60–85 years.

The primary hypothesis is that the ACTIVE group will have a higher physical function composite Z-score at week 8 than the control group. Other measures of physical function, physical activity and health-related quality of life will be evaluated as secondary outcomes.

## Methods

### Study design

This is a single-centre, assessor-blinded, two-arm, parallel-group randomised controlled trial. The protocol was prepared in accordance with the SPIRIT 2025 statement.^19^ The completed checklist is provided in online supplemental file 1. The term “community-dwelling” refers to the participants’ place of residence. All participants live independently in the community, and hospital outpatient clinics are one of several recruitment channels.

### Study setting

The investigators will recruit 142 participants through Peking University Third Hospital and in Haidian District and neighbouring communities in Beijing. Recruitment was scheduled to start in March 2026, with data collection expected to be completed in December 2026 (figure 1).

**Figure 1.**
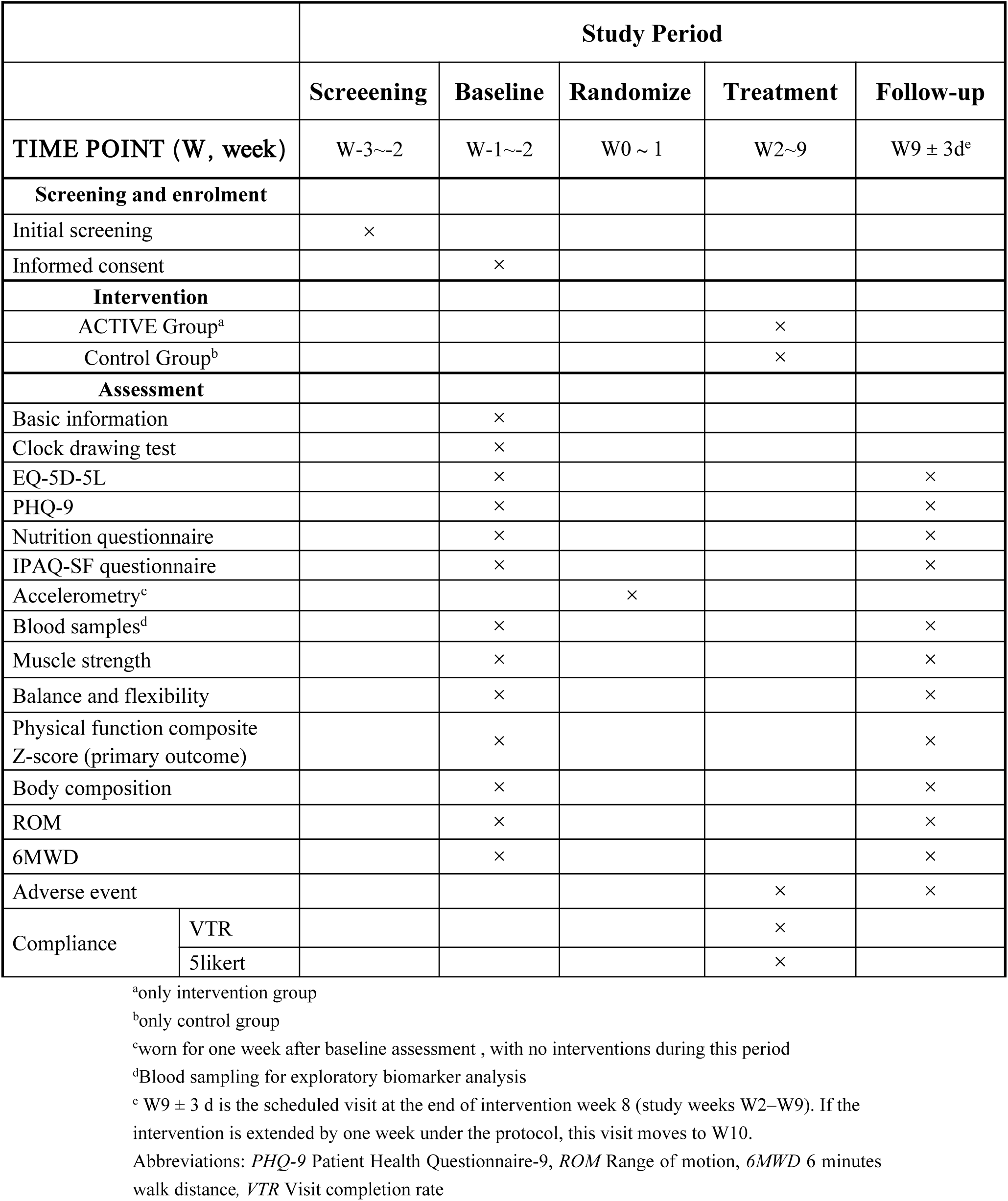
Trial schedule. Timing of screening, baseline assessment, randomisation, the 8-week intervention and follow-up assessment.

### Eligibility criteria

#### Inclusion criteria

- Aged 60–85 years.
- No major clinical condition requiring treatment.
- Willingness to provide written informed consent.

#### Exclusion criteria

- Heart failure of Killip class II or higher (eg, chest pain when walking 100 or 200 m or climbing one flight of stairs).
- Major cardiac surgery in the previous 6 months, including valve replacement or bypass surgery.
- Stroke in the previous 12 months.
- Parkinson’s disease or diagnosed dementia.
- A fracture or fragility fracture in the previous year.
- Severe kidney disease requiring dialysis.
- Inability to complete basic functional tests, such as the SPPB or the single-leg stance test.
- Arthritis severe enough to impede physical activity.
- Deemed unsuitable for participation by the investigators.

### Screening and randomisation

Participants will be recruited through outpatient clinics at Peking University Third Hospital, community posters and media advertisements.

Eligible participants will provide written informed consent. A web-based central randomisation system implemented in REDCap will use simple randomisation to assign participants 1:1 to the ACTIVE group or the standardised, non-individualised exercise group (figure 2). Allocation will remain concealed until the system assigns the participant. Each participant will receive a unique study identification number.

**Figure 2.**
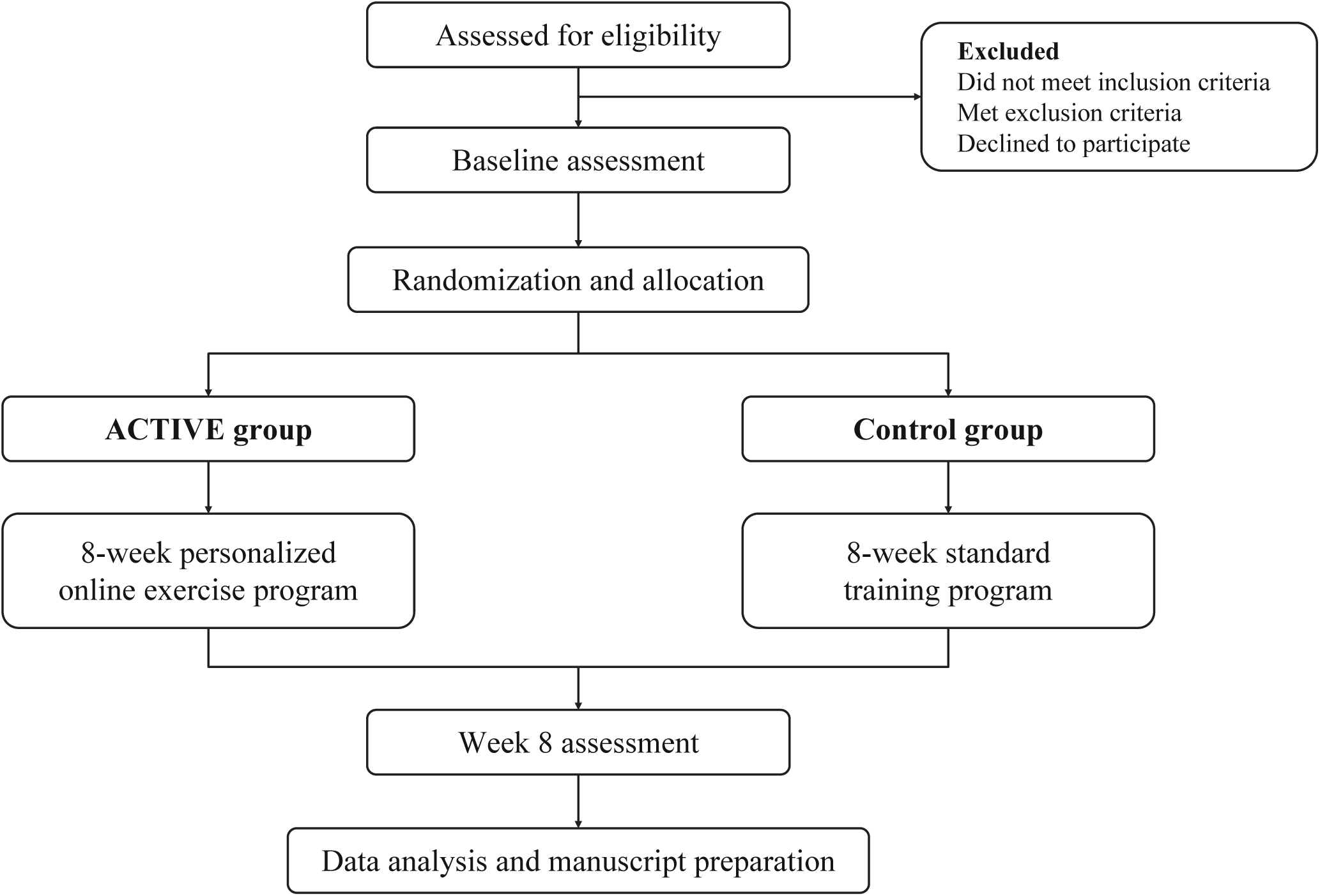
Trial flow diagram. Participant recruitment, screening, enrolment, randomisation, intervention completion and follow-up. For participants whose intervention is extended by one week under the protocol, the end-of-intervention assessment is postponed accordingly.

### Blinding

Given the nature of the exercise intervention, blinding participants and those delivering the intervention is infeasible. Outcome assessors and data analysts will remain blinded to group allocation throughout the study. No unblinding of outcome assessors or data analysts is planned during the trial.

### Interventions

Both groups will receive their intervention through a WeChat mini programme. Specialists in sports medicine and rehabilitation, a medical device and information technology development team, and a professional video production team developed the mini programme over 9 months.

The mini programme has participant and clinician interfaces (figure 3). Participants can access exercise, health education, consultation and check-in functions. Clinicians can monitor exercise completion in real time, receive enquiries and provide timely feedback.

**Figure 3.**
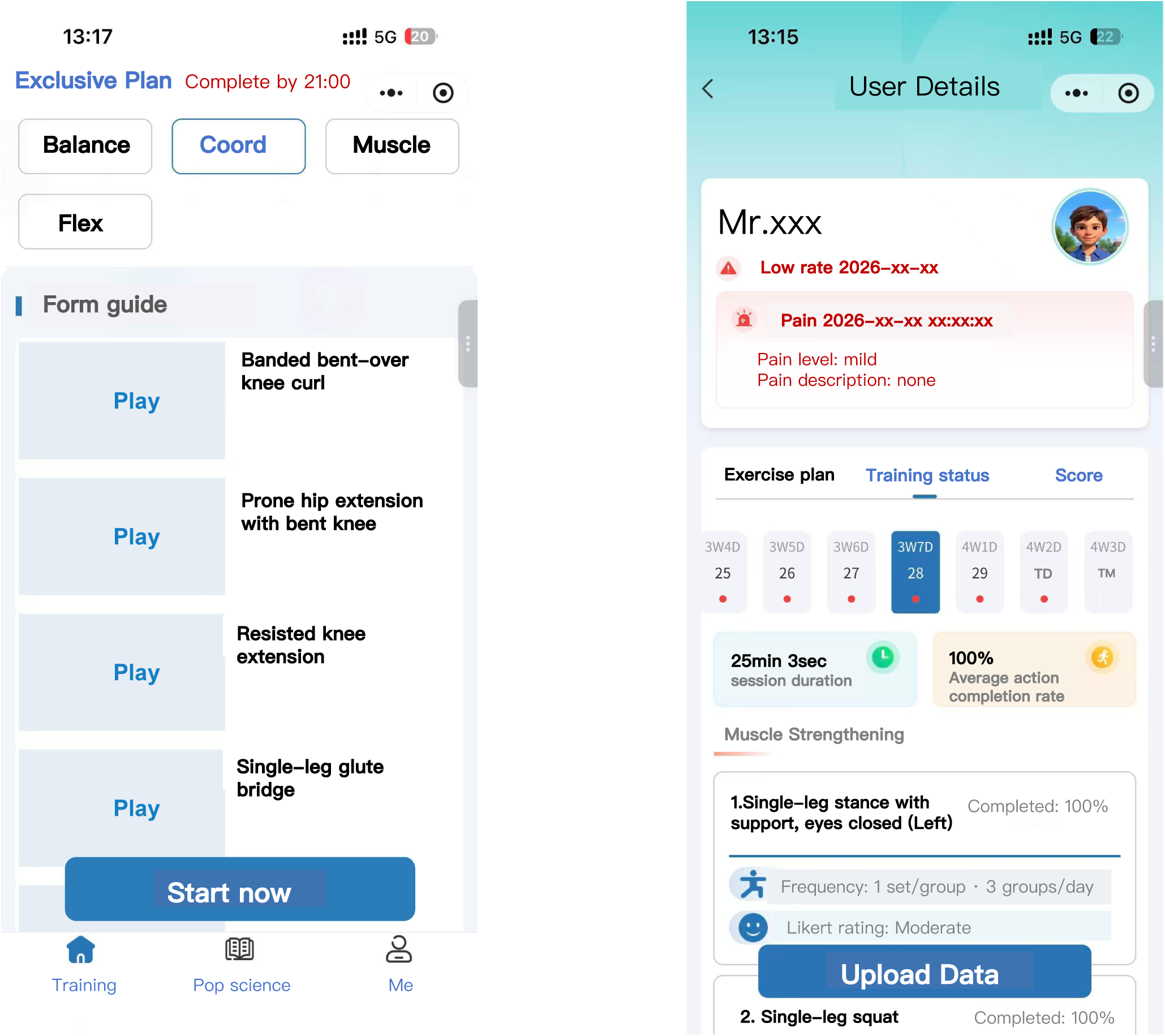
Participant and clinician interfaces of the WeChat mini programme. The participant interface (left) shows the exercise plan and instructional videos. The clinician interface (right) shows session duration, exercise completion rates, participant-reported difficulty ratings, and alerts for low completion or reported pain.

### ACTIVE intervention group

Participants in the ACTIVE group will receive an individualised home-based exercise programme generated by the rule-based exercise prescription system and delivered through the WeChat mini programme. The core decision rules were established before the trial. The system uses fixed rules, with no learning from online data or updating of the rules during the trial.

The system applies prespecified thresholds to convert assessment results into structured labels. These labels guide identification of contraindications, selection of exercise modules, setting of the starting dose and adjustment of difficulty. Online supplemental file 2 provides the thresholds and decision rules, together with their literature or expert basis.

The system selects appropriate content from a library of 131 prerecorded exercise videos to generate each exercise prescription. It then adjusts the difficulty in response to training feedback submitted by participants.

All participants in the ACTIVE group will receive one-to-one instruction from a rehabilitation therapist at the outpatient clinic on how to perform the exercises and use the WeChat mini programme. They will then exercise at home according to the prescribed frequency, session duration and combination of modules. After each module, participants will rate perceived difficulty on a five-point scale (1=very easy, 2=easy, 3=moderate, 4=difficult and 5=very difficult). If the same module is rated easy on two consecutive occasions or very easy once, the system will automatically increase its difficulty. When a module is too difficult, the system will reduce its difficulty according to the prespecified rules.

### Standardised exercise control group

The control group will receive an 8-week standardised, non-individualised home-based exercise programme through the same WeChat mini programme. The programme uses a fixed set of 12 distinct exercises from the exercise library covering strength, balance, coordination and flexibility exercise. Baseline functional assessments do not determine exercise content in this group.

Control group participants will also receive initial one-to-one instruction at the outpatient clinic on the exercises and use of the WeChat mini programme, then follow the prespecified standardised programme at home. The mini programme will provide reminders, monitor participation and record daily video completion rates in this group. The programme does not use participant feedback to progress or regress exercise difficulty in the control group.

### Shared intervention components and delivery

Both groups will undertake 8 weeks of home-based exercise through the same WeChat mini programme, with the same daily reminders, initial one-to-one in-person instruction, health education, monitoring and feedback procedures. In the ACTIVE group, daily training duration will depend on baseline functional level and may be adjusted in response to training feedback. The control group will follow a prespecified schedule. The platform, intervention period, reminder frequency, and procedures for monitoring and feedback will be the same in both groups. The system will record training data for both groups. Routine clinical care is permitted. Participants must not enrol in another exercise rehabilitation study during the trial and must inform the study physician before starting any new intervention.

### Exercise adherence

The WeChat mini programme will send a reminder at 06:00 each day and automatically record each participant’s daily video completion rate by module and their check-in frequency. If the daily completion rate falls below 60%, the clinician interface will generate an alert and the study team will follow up by telephone to support adherence. If the average completion rate falls below 30% in any week of the 8-week intervention, the intervention will be extended by 1 week and the follow-up assessment will be postponed accordingly.

### Outcomes

#### Assessment schedule and data collection

Primary and secondary efficacy outcomes will be measured at the baseline visit before randomisation and at the end-of-intervention visit, scheduled for week 8. The final assessment will be postponed accordingly for participants whose intervention is extended under the protocol. Before the trial begins, assessors blinded to group allocation will receive standardised training in the assessment procedures. Peking University Clinical Research Institute will perform periodic checks.

#### Primary outcome

The primary outcome is the physical function composite Z-score at week 8, comprising 4-m gait speed, five-times sit-to-stand time, 6-min walk distance and eyes-open single-leg stance time. This composite was selected because the SPPB total score may have a ceiling effect in older adults with relatively preserved function.^20,21^ Following the method for aggregating standardised measures used in the CardioRACE trial,^22^ the mean and SD for each component will be calculated using all available baseline data from randomised participants. The same parameters will be used at baseline and at the final visit to calculate each component Z-score as (individual measurement − overall baseline mean) ÷ overall baseline SD. The sign of the five-times sit-to-stand Z-score will be reversed so that higher scores indicate better function for all components. The composite will be calculated as the unweighted arithmetic mean of the four Z-scores when all four measurements are valid at the same visit.

The component measures will be obtained as follows.

##### 4-m gait speed and 6-min walk distance

Participants will walk back and forth along a marked course, covering as much distance as possible in 6 min. They will refrain from running, jumping and talking and may stop to rest and resume walking if needed. Assessors will use a stopwatch to record the time taken to reach the 4-m mark. Gait speed (m/s) will be calculated by dividing 4 m by this time. During the test, assessors will announce the time and provide standardised encouragement each minute. They will give a warning 15 s before the end, mark the stopping position and record the total distance walked in 6 min (m).^23^

##### Five-times sit-to-stand test

A 43-cm-high chair with a backrest and no armrests will be used. Participants will sit with their knees shoulder-width apart and their arms crossed over their chest. On command, they will complete five sit-to-stand repetitions as quickly as possible without using their arms for support. Assessors will record the completion time in seconds using a stopwatch.

##### Eyes-open single-leg stance test

With their hands on their hips and their gaze forward, participants will lift the non-supporting foot approximately 10 cm off the floor and maintain balance for as long as possible. Assessors will stop timing when the raised foot touches the floor. Each side will be tested twice, with the longer duration in seconds retained for each side. The arithmetic mean of the left and right values will be used as the single-leg stance component of the composite score.^24^

#### Secondary outcomes

##### Physical function components

The individual Z-scores and raw values of the four component measures will be analysed separately. Raw eyes-open single-leg stance times will be reported for the left side, the right side and the side with the shorter stance time at baseline. The latter side will remain fixed for the final assessment.

##### Physical activity

The IPAQ-SF^25,26^ will be scored according to the standard scoring guidelines.^27^

##### Health-related quality of life

The EQ-5D-5L^28^ will be scored using the Chinese value set developed by Luo and colleagues^29^ to derive the health utility value for analysis. Participants will also verbally report a rating of their overall health.

##### Flexibility

Flexibility will be assessed using the chair sit-and-reach test.^30^

##### Lower-limb muscle strength and asymmetry

A microFET3 handheld dynamometer and a stabilisation belt will be used to measure maximal isometric force bilaterally in the gluteus medius, gluteus maximus, hip adductors, quadriceps and hamstrings.^31^ The lower-limb strength asymmetry index (ASI) will be calculated from the bilateral measurements obtained at the same visit. The hamstrings-to-quadriceps (H:Q) ratio will be calculated by dividing peak hamstring force by peak quadriceps force on the same side.

##### Handgrip strength

Handgrip strength will be measured with a Jamar dynamometer. The higher of the maximum values for the two hands (kg) will be used in the analysis.^32^

#### Safety outcomes

Safety outcomes comprise adverse events (AEs), serious adverse events (SAEs), adverse reactions (ARs) and serious adverse reactions (SAEs considered at least possibly related to the intervention) occurring during the study.

### Sample size

The sample size was estimated using the standardised between-group mean difference in SPPB scores reported in previous research. In the Vivifrail randomised trial, Casas-Herrero and colleagues reported between-group differences in SPPB score change of 0.86 points at 1 month and 1.40 points at 3 months for the intervention versus usual care.^11^ Standard errors were approximated from the 95% CIs for the changes in each group, and SDs were then estimated using the numbers randomised to each group. Dividing each between-group difference by the pooled SD yielded approximate reference effect sizes (Cohen’s d) of 0.46 and 0.70, respectively. The present trial uses d=0.50 as the design assumption, given that the earlier estimates came from linear mixed models and that the population, comparator and outcome measure differed from those in this trial.

The required sample size was calculated using G*Power (V.3.1.9.7).^33^ A two-sided independent-samples t-test with a 1:1 allocation ratio, a significance level of α=0.05, 80% power and d=0.50 required 64 analysable participants per group, or 128 in total. Allowing for 10% attrition, the target is 142 randomised participants, with 71 per group. This calculation did not account for any gain in precision from baseline adjustment. The primary outcome will be analysed using baseline-adjusted analysis of covariance (ANCOVA), as specified in the statistical analysis plan.

### Statistical analysis

Analyses will follow the statistical analysis plan finalised on 23 October 2026. Efficacy analyses will use both the full analysis set (FAS) and the per-protocol set (PPS), with the FAS as the primary analysis population. The FAS includes all randomised participants except those who fail to meet the eligibility criteria or withdraw after randomisation before receiving any intervention. Participants will be analysed in their assigned groups for efficacy. The PPS includes eligible participants who received the allocated intervention according to the protocol without major deviations, completed the final visit within the specified window and have complete baseline and final measurements for all four components of the primary outcome. Major deviations, including fewer than 32 days of training, will result in exclusion from the PPS. The safety analysis set includes participants who received at least one intervention session after randomisation and have safety data. Safety analyses will be conducted according to the intervention received.

Baseline characteristics will be summarised primarily using descriptive statistics, without formal between-group hypothesis tests. The primary outcome will be analysed using ANCOVA with the week 8 physical function composite Z-score as the dependent variable, randomised group as a fixed effect and the baseline composite Z-score as a covariate. The adjusted between-group difference, two-sided 95% CI and p value will be reported. The primary analysis will use observed data from FAS participants with primary outcome measurements at baseline and week 8. No single imputation will be used. The proportion, reasons and patterns of missing data will be summarised by randomised group, visit and outcome measure. Sensitivity analyses will be guided by the missing data. Where necessary, multiple imputation will be used under the missing-at-random assumption. Robustness will be assessed under different assumptions about the missingness mechanism.

Sensitivity analyses of the primary outcome will repeat the primary analysis in the PPS. If important baseline characteristics such as age, sex, body mass index (BMI) or comorbidities show substantial between-group imbalance, the ANCOVA model will additionally adjust for those variables. Continuous secondary outcomes will be analysed using ANCOVA with the week 8 value as the dependent variable, group as a fixed effect and the corresponding baseline value as a covariate. Adjusted between-group mean differences, 95% CIs and p values will be reported. For categorical outcomes, counts (n/N) and proportions will be reported by group, with between-group effect estimates and 95% CIs obtained using logistic regression. The proportions of participants with AEs or SAEs will be compared between groups using the χZ test or Fisher’s exact test. Statistical analyses will be performed using SAS V.9.4. All tests will be two-sided, and p≤0.05 will indicate statistical significance. No interim analyses are planned.

### Data management

#### Data security and storage

REDCap is hosted on Alibaba Cloud servers. All data transmissions are protected by end-to-end TLS 1.3 encryption. Role-based access controls will assign permissions according to study roles (PI, CRC, CRA and PM), and a complete audit trail of all system activity will be retained.

The WeChat mini programme will record video playback completion rates for each exercise module, check-ins, participant-reported difficulty ratings and system records of progression or regression. These data will be synchronised with the backend database. Automatic recording reduces manual transcription and provides a continuous record of platform use and exercise completion.

Paper source documents, including signed consent forms, assessment checklists and exercise video review records, will be kept in sealed archives at the Institute of Sports Medicine, Peking University Third Hospital, for at least 10 years. After trial completion, all electronic data will be encrypted and stored on institutional servers for the same period.

### Ethics and dissemination

#### Dissemination

Trial results will be published in a peer-reviewed journal.

#### Participant safety

Before starting the intervention, all participants will undergo a comprehensive risk assessment covering health and exercise function. Adverse events will be collected, documented and reported throughout the intervention.

#### Adverse events

An AE is any untoward medical occurrence during the study, including a new illness, worsening of pre-existing symptoms or exercise-related discomfort. Expected responses to exercise, such as muscle soreness that resolves within 48 hours, are excluded. An SAE is an event that results in death, is life-threatening, leads to hospitalisation or permanent functional impairment, or requires urgent medical intervention.

The relationship of each AE to the intervention will be rated as definite, probable, possible, unlikely or unrelated. Events in the first three categories will be classified as ARs. Any AE during the intervention will trigger suspension of exercise and an alert in the clinician interface. Participants may resume exercise only after review and clearance by a clinician (online supplemental file 2, table S7). Each AE will be managed promptly according to its severity. Non-serious events will be treated symptomatically, with escalation if symptoms persist. SAEs will trigger immediate emergency management and referral. Full details of all AEs will be documented within 24 hours. SAEs will also be reported in writing to the ethics committee and the sponsor within 24 hours, followed by further follow-up. An event will be classified as resolved if symptoms disappear, sequelae stabilise or the participant is lost to follow-up.

### Trial discontinuation and participant withdrawal

The investigators may discontinue the study or require a participant to withdraw in the following circumstances.

- The participant needs to withdraw because of relocation or personal health reasons, including an acute illness, an SAE or an intolerable adverse reaction such as exercise-induced angina, syncope or fracture.
- The participant’s completion rate is below 30% for two consecutive weeks, making adequate exposure to the intervention unlikely.

Participants may withdraw voluntarily. When a participant requests withdrawal, the study coordinator will confirm the decision, discuss and document the main reason, such as personal reasons, health problems or dissatisfaction with the intervention. All data collected before withdrawal will be retained in the analysis dataset.

Regardless of the reason for withdrawal, participants will be contacted for safety follow-up, usually within 1–2 weeks of the decision to withdraw, to establish their health status and document any AEs potentially related to the intervention.

The trial may be stopped early for serious safety concerns, major protocol flaws or deviations that prevent evaluation of the intervention effect, or at the request of the trial applicant or a regulatory authority. If early termination is necessary, the investigators will inform participants and report it to the ethics committee.

### Trial monitoring

Peking University Clinical Research Institute (PUCRI) has been commissioned to monitor the trial. PUCRI will conduct regular on-site monitoring visits in accordance with the monitoring plan to safeguard participants’ rights and safety, verify the integrity and reliability of trial data, and ensure that the trial is conducted in accordance with applicable regulations and the protocol.

### Equity, diversity and inclusion (PRO EDI)

The trial incorporates several measures to promote equity, diversity and inclusion.

To address potential barriers related to limited digital literacy,^34^ all participants will be shown how to use the WeChat mini programme before their first exercise session. They can raise health or technical concerns directly through the consultation function and receive a response from a rehabilitation therapist.

Recruitment through both hospital outpatient clinics and community channels is intended to reach people from a range of socioeconomic and educational backgrounds. The broad age range (60–85 years) and eligibility regardless of sex further support sample diversity.

The mini programme interface was designed with input from five older adults aged 65–80 years on the clarity of exercise instructions and interface usability.

## Discussion

The ACTIVE trial asks whether individualised exercise prescriptions informed by multidimensional baseline assessment and training feedback provide greater functional benefits than standardised, non-individualised multicomponent exercise when both groups receive the same digital platform, exercise reminders, initial professional instruction, monitoring and feedback. In a home-based exercise study, Dondzila and colleagues found that a programme combining individualised step goals and resistance band training improved some functional outcomes.^35^ Because both goal setting and exercise components differed between groups, the study could not isolate the effect of individualised prescription. The trials by Li and colleagues and the StandingTall investigators also support using digital platforms to deliver home-based exercise.^15,16^ Li and colleagues used health education as the comparator, while StandingTall focused primarily on balance training. Neither compared individualised and non-individualised exercise on the same digital platform with a common multicomponent exercise framework. In the present trial, both groups may benefit because the active comparator is standardised, non-individualised multicomponent exercise. By providing the same platform and support to both groups, the trial will estimate the added effect of the individualised prescription strategy.

We expect the ACTIVE group to have a higher physical function composite Z-score at the end of the intervention than the control group. The expected benefit may arise in two ways. First, baseline assessment can identify participants with generally good function who nevertheless have specific limitations, such as poorer balance on one side or weakness in particular lower-limb muscle groups, and prioritise exercises for the relevant function or side. Second, progressing or regressing each component within safety limits can help participants with lower initial function start with exercises they can perform and continue to challenge those who adapt more quickly. This design translates the functional assessment and progressive loading principles in exercise recommendations,^7,8^ together with the use of functional stratification and perceived exertion feedback in earlier programmes,^11,15^ into explicit rules for module selection, starting dose and difficulty adjustment. Exercise completion, cumulative training duration and AEs will also be assessed to determine whether between-group differences in function are accompanied by better programme adherence or tolerability.

The primary outcome combines gait speed, sit-to-stand performance, walking endurance and static balance in a composite Z-score. It is intended to capture the overall effects of multicomponent exercise in community-dwelling older adults with relatively preserved function and to limit the influence of the potential ceiling effect of the SPPB total score on interpretation.^20,21^ The method for aggregating standardised scores was based on the CardioRACE trial.^22^ The clinical significance of changes in this particular composite remains to be established. The individual components will help identify the functional domains in which the groups differ. Of the four components, the five-times sit-to-stand test and eyes-open single-leg stance time are most directly aligned with ACTIVE exercises. In participants with slower sit-to-stand performance or lower limb strength at baseline, progressive resistance exercise targeting the gluteal and leg muscles may shorten sit-to-stand time by improving force production and movement control. For participants with a shorter stance time on one side at baseline, side-specific balance training may first improve performance on that side. Among the remaining secondary outcomes, changes are more likely in the muscle groups, sides or ranges of motion targeted by training, such as strength on the weaker side or chair sit-and-reach performance in participants with limited flexibility. Bilateral raw strength measurements should be considered when interpreting the ASI to determine whether a smaller difference reflects gains on the weaker side. The IPAQ-SF assesses self-reported physical activity. Completing the prescribed exercise may leave overall daily activity unchanged. The EQ-5D-5L covers several dimensions of health, and a substantial proportion of respondents in a study of community-dwelling older adults in China reported full health.^36^ Thus, objectively measured function may improve in the short term even if between-group differences in physical activity or health utility are small.

If the primary outcome favours ACTIVE and the direction of effects on sit-to-stand performance, balance or relevant strength measures is consistent with the training content, these results will help identify the domains that benefit. If the composite score shows no clear between-group difference, the precision of the effect estimate and the direction of effects on its components should be considered when judging whether the findings remain compatible with a small benefit. If adherence is good in both groups and the effect estimate is close to zero, the standardised programme may already meet the short-term exercise needs of most participants.

The findings will primarily inform assessment of the short-term effects of ACTIVE in independently living older adults in urban Beijing. Multicentre validation will follow. Regardless of whether ACTIVE provides additional functional benefits, this trial will provide evidence on the value of multidimensional baseline assessment and feedback-based adjustments in community exercise prescription and inform future programme refinement and implementation research.

## Supporting information

SPIRIT2025 checklist

## Trial status

This is protocol V.5.0, dated 9 July 2026. Recruitment was scheduled to start in March 2026, and data collection is expected to be completed in December 2026. Recruitment had been completed at the time of manuscript submission.

## Abbreviations

ACTIVE: knowledge-based, rule-driven exercise prescription decision system
AE: adverse event
ANCOVA: analysis of covariance
AR: adverse reaction
ASI: strength asymmetry index
ASMI: appendicular skeletal muscle mass index
AWGS: Asian Working Group for Sarcopenia
CDT: clock drawing test
DMP: data management plan
DSMB: data and safety monitoring board
EQ-5D-5L: EuroQol five-dimension, five-level questionnaire
H:Q ratio: hamstrings-to-quadriceps ratio
IPAQ-SF: International Physical Activity Questionnaire Short Form
ITT: intention to treat
MVPA: moderate-to-vigorous physical activity
PHQ-9: Patient Health Questionnaire-9
PUCRI: Peking University Clinical Research Institute
RCT: randomised controlled trial
REDCap: Research Electronic Data Capture
SAE: serious adverse event
SPIRIT: Standard Protocol Items: Recommendations for Interventional Trials
SPPB: Short Physical Performance Battery
SSL: Secure Sockets Layer

## Declarations

### Ethics approval and consent to participate

This study was approved by the Medical Science Research Ethics Committee of Peking University Third Hospital (approval no. (2025) 991-01). All participants will provide written informed consent before any study procedures. Protocol amendments will be submitted to the ethics committee for approval before implementation. Written consent for trial participation covers blood collection at baseline and at the end-of-intervention visit for exploratory biomarker analyses within this trial. Blood specimens will be disposed of according to routine clinical procedures after testing.

### Consent for publication

Not applicable, as this manuscript contains no individual participant data. The informed consent form template is available on request.

### Data availability statement

The principal investigators (YX, XM and ZW), co-investigators (YP, HL, YL and YH) and trial monitors (PUCRI) will have access to the final trial dataset. No contractual agreement restricts their access. To protect participants’ privacy, the datasets generated and analysed in this study will be kept private and will be available from the corresponding authors on reasonable request.

## Trial sponsor and funder roles

The trial sponsor is Peking University Third Hospital. The contact for sponsor enquiries is Yan Xu, Institute of Sports Medicine, Peking University Third Hospital, 49 North Garden Road, Haidian District, Beijing 100191, China.

The study investigators designed the trial and are responsible for its conduct, data analysis and reporting. Peking University Third Hospital is the trial site, receives serious adverse event reports and retains institutional approval and decision-making authority for the trial. The grant programmes listed under Funding provide financial support only.

## Contributors

YP, HL, YL, YH and MQ conceived the study and designed the protocol. YP drafted the manuscript. ZW, XM and YX supervised the project and secured funding. All authors reviewed the manuscript and approved the final version.

## Acknowledgements

We thank all older participants for their time and effort. We also thank the research team at the Institute of Sports Medicine, Peking University Third Hospital, and Peking University Clinical Research Institute for their clinical and technical support.

## Online supplemental files

**Online supplemental file 1.** Completed SPIRIT 2025 checklist, with page numbers for the corresponding items in the protocol.

**Online supplemental file 2.** ACTIVE decision rules, including prespecified clinical thresholds for assigning structured labels, the exercise module selection algorithm, the starting-dose table and the rules for progression and regression.

## References

1 National Bureau of Statistics of China, Office of the Leading Group of the State Council for the Seventh National Population Census. Communiqué of the Seventh National Population Census (No. 5): age composition. 11 May 2021. Available: https://www.stats.gov.cn/english/PressRelease/202105/t20210510_1817190.html

2 World Health Organization. Healthy ageing and functional ability. 2020. Available: https://www.who.int/philippines/news/q-a-detail/healthy-ageing-and-functional-ability [Accessed 2 Nov 2026].

3 Guralnik JM, Ferrucci L, Simonsick EM, et al. Lower-extremity function in persons over the age of 70 years as a predictor of subsequent disability. N Engl J Med 1995;332:556–61. doi:10.1056/NEJM199503023320902

4 Yu B, Zhou Z, Chen G, et al. Intrinsic capacity subtypes in healthy Chinese older adults: links to possible sarcopenia and fall risk. J Nutr Health Aging 2026;30:100931. doi:10.1016/j.jnha.2026.100931

5 Nursing Group of Chinese Society of Geriatrics, Committee on Geriatric Nursing of Chinese Association of Geriatric Research, China Gerontological Nursing Alliance, et al. Clinical practice guidelines for exercise interventions for the prevention of disability in older adults (2023). Chinese General Practice 2023;26:2695–710. doi:10.12114/j.issn.1007-9572.2023.0223

6 Snowsill TM, Stathi A, Green C, et al. Cost-effectiveness of a physical activity and behaviour maintenance programme on functional mobility decline in older adults: an economic evaluation of the REACT (Retirement in Action) trial. Lancet Public Health 2022;7:e327–34. doi:10.1016/S2468-2667(22)00030-5

7 Izquierdo M, Merchant RA, Morley JE, et al. International exercise recommendations in older adults (ICFSR): expert consensus guidelines. J Nutr Health Aging 2021;25:824–53. doi:10.1007/s12603-021-1665-8

8 Nelson ME, Rejeski WJ, Blair SN, et al. Physical activity and public health in older adults: recommendation from the American College of Sports Medicine and the American Heart Association. Med Sci Sports Exerc 2007;39:1435–45. doi:10.1249/mss.0b013e3180616aa2

9 Stathi A, Greaves CJ, Thompson JL, et al. Effect of a physical activity and behaviour maintenance programme on functional mobility decline in older adults: the REACT (Retirement in Action) randomised controlled trial. Lancet Public Health 2022;7:e316–26. doi:10.1016/S2468-2667(22)00004-4

10 Pahor M, Guralnik JM, Ambrosius WT, et al. Effect of structured physical activity on prevention of major mobility disability in older adults: the LIFE study randomized clinical trial. JAMA 2014;311:2387–96. doi:10.1001/jama.2014.5616

11 Casas-Herrero Á, Sáez de Asteasu ML, Antón-Rodrigo I, et al. Effects of Vivifrail multicomponent intervention on functional capacity: a multicentre, randomized controlled trial. J Cachexia Sarcopenia Muscle 2022;13:884–93. doi:10.1002/jcsm.12925

12 Chen B, Li M, Zhao H, et al. Effect of multicomponent intervention on functional decline in Chinese older adults: a multicenter randomized clinical trial. J Nutr Health Aging 2023;27:1063–75. doi:10.1007/s12603-023-2031-9

13 Pedersen BK, Saltin B. Exercise as medicine: evidence for prescribing exercise as therapy in 26 different chronic diseases. Scand J Med Sci Sports 2015;25 Suppl 3:1–72. doi:10.1111/sms.12581

14 Fiuza-Luces C, Garatachea N, Berger NA, et al. Exercise is the real polypill. Physiology (Bethesda) 2013;28:330–58. doi:10.1152/physiol.00019.2013

15 Delbaere K, Valenzuela T, Lord SR, et al. E-health StandingTall balance exercise for fall prevention in older people: results of a two year randomised controlled trial. BMJ 2021;373:n740. doi:10.1136/bmj.n740

16 Li N, Wang N, Xu Y, et al. The impacts of a mHealth platform-enabled lifestyle-integrated multicomponent exercise program on reversing pre-frailty in community-dwelling older adults: a randomized controlled trial. Int J Nurs Stud 2025;167:105072. doi:10.1016/j.ijnurstu.2025.105072

17 World Health Organization. WHO’s work on the UN Decade of Healthy Ageing (2021–2030). Available: https://www.who.int/initiatives/decade-of-healthy-ageing

18 Fang EF, Xie C, Schenkel JA, et al. A research agenda for ageing in China in the 21st century (2nd edition): focusing on basic and translational research, long-term care, policy and social networks. Ageing Res Rev 2020;64:101174. doi:10.1016/j.arr.2020.101174

19 Chan AW, Boutron I, Hopewell S, et al. SPIRIT 2025 statement: updated guideline for protocols of randomised trials. BMJ 2025;389:e081477. doi:10.1136/bmj-2024-081477

20 Lamb SE, Keene DJ. Measuring physical capacity and performance in older people. Best Pract Res Clin Rheumatol 2017;31:243–54. doi:10.1016/j.berh.2017.11.008

21 Pua YH, Tay L, Lin S, et al. Validity and utility of a modified Short Physical Performance Battery assessment and scoring approach in middle-aged and older adults. J Am Med Dir Assoc 2026;27:106027. doi:10.1016/j.jamda.2025.106027

22 Lee DC, Brellenthin AG, Lanningham-Foster LM, et al. Aerobic, resistance, or combined exercise training and cardiovascular risk profile in overweight or obese adults: the CardioRACE trial. Eur Heart J 2024;45:1127–42. doi:10.1093/eurheartj/ehad827

23 ATS Committee on Proficiency Standards for Clinical Pulmonary Function Laboratories. ATS statement: guidelines for the six-minute walk test. Am J Respir Crit Care Med 2002;166:111–7. doi:10.1164/ajrccm.166.1.at1102

24 Springer BA, Marin R, Cyhan T, et al. Normative values for the unipedal stance test with eyes open and closed. J Geriatr Phys Ther 2007;30:8–15. doi:10.1519/00139143-200704000-00003

25 Craig CL, Marshall AL, Sjöström M, et al. International physical activity questionnaire: 12-country reliability and validity. Med Sci Sports Exerc 2003;35:1381–95. doi:10.1249/01.MSS.0000078924.61453.FB

26 Deng HB, Macfarlane DJ, Thomas GN, et al. Reliability and validity of the IPAQ-Chinese: the Guangzhou Biobank Cohort study. Med Sci Sports Exerc 2008;40:303–7. doi:10.1249/mss.0b013e31815b0db5

27 IPAQ Research Committee. Guidelines for data processing and analysis of the International Physical Activity Questionnaire (IPAQ): short and long forms. November 2005. Available: https://biobank.ndph.ox.ac.uk/ukb/ukb/docs/ipaq_analysis.pdf

28 Herdman M, Gudex C, Lloyd A, et al. Development and preliminary testing of the new five-level version of EQ-5D (EQ-5D-5L). Qual Life Res 2011;20:1727–36. doi:10.1007/s11136-011-9903-x

29 Luo N, Liu G, Li M, et al. Estimating an EQ-5D-5L value set for China. Value Health 2017;20:662–9. doi:10.1016/j.jval.2016.11.016

30 Jones CJ, Rikli RE, Max J, et al. The reliability and validity of a chair sit-and-reach test as a measure of hamstring flexibility in older adults. Res Q Exerc Sport 1998;69:338–43. doi:10.1080/02701367.1998.10607708

31 Grootswagers P, Vaes AMM, Hangelbroek R, et al. Relative validity and reliability of isometric lower extremity strength assessment in older adults by using a handheld dynamometer. Sports Health 2022;14:899–905. doi:10.1177/19417381211063847

32 Roberts HC, Denison HJ, Martin HJ, et al. A review of the measurement of grip strength in clinical and epidemiological studies: towards a standardised approach. Age Ageing 2011;40:423–9. doi:10.1093/ageing/afr051

33 Faul F, Erdfelder E, Lang AG, et al. G*Power 3: a flexible statistical power analysis program for the social, behavioral, and biomedical sciences. Behav Res Methods 2007;39:175–91. doi:10.3758/BF03193146

34 Liu L, Wu F, Tong H, et al. The digital divide and active aging in China. Int J Environ Res Public Health 2021;18:12675. doi:10.3390/ijerph182312675

35 Dondzila CJ, Swartz AM, Keenan KG, et al. Translating exercise interventions to an in-home setting for seniors: preliminary impact on physical activity and function. Aging Clin Exp Res 2016;28:1227–35. doi:10.1007/s40520-015-0518-x

36 You R, Liu J, Yang Z, et al. Comparing the performance of the EQ-5D-3L and the EQ-5D-5L in an elderly Chinese population. Health Qual Life Outcomes 2020;18:97. doi:10.1186/s12955-020-01324-0

