## Supplementary material for "Effects of an exercise prescription system (ACTIVE) on physical function in community-dwelling older adults: protocol for a single-centre, assessor-blinded, parallel-group randomised controlled trial": SPIRIT2025 checklist

### SPIRIT 2025 checklist of items to address in a randomized trial protocol*

Instructions: Complete this checklist by entering the page numbers from your manuscript where readers will find each item. If an item does not apply, write "N/A" and provide a brief explanation.

Citation: Chan A-W, Boutron I, Hopewell S, Moher D, Schulz KF, et al. SPIRIT 2025 statement: updated guideline for protocols of randomised trials. BMJ 2025;389:e081477.

| Section / Topic | No | SPIRIT 2025 Checklist Item | Page |
|---|---|---|---|
| **Administrative information** | | | |
| Title and structured summary | 1a | Title stating the trial design, population, and interventions, with identification as a protocol | 1 |
| | 1b | Structured summary of trial design and methods, including items from the World Health Organization Trial Registration Data Set | 2 (selected WHO registration items) |
| Protocol version | 2 | Version date and identifier | 16 |
| Roles and responsibilities | 3a | Names, affiliations, and roles of protocol contributors | 1, 19 |
| | 3b | Name and contact information for the trial sponsor | 19 |
| | 3c | Role of trial sponsor and funders in design, conduct, analysis, and reporting of trial; including any authority over these activities | 19 |
| | 3d | Composition, roles, and responsibilities of the coordinating site, steering committee, endpoint adjudication committee, data management team, and other individuals or groups overseeing the trial, if applicable | 12, 14, 19 (oversight roles partly described) |
| **Open science** | | | |
| Trial registration | 4 | Name of trial registry, identifying number (with URL), and date of registration. If not yet registered, name of intended registry | 2 |
| Protocol and statistical analysis plan | 5 | Where the trial protocol and statistical analysis plan can be accessed | 11, 16 (access arrangements not reported) |
| Data sharing | 6 | Where and how the individual de-identified participant data (including data dictionary), statistical code, and any other materials will be accessible | 18 (dataset access described; code and data dictionary not specified) |
| Funding and conflicts of interest | 7a | Sources of funding and other support (e.g., supply of drugs) | 19 |
| | 7b | Financial and other conflicts of interest for principal investigators and steering committee members | 18 |
| Dissemination policy | 8 | Plans to communicate trial results to participants, healthcare professionals, the public, and other relevant groups (e.g., reporting in trial registry, plain language summary, publication) | 12 |
| **Introduction** | | | |
| Background and rationale | 9a | Scientific background and rationale, including summary of relevant studies (published and | 3–5 (benefits discussed; harms evidence limited) |

| | | | |
|---|---|---|---|
| | | unpublished) examining benefits and harms for each intervention | |
| | 9b | Explanation for choice of comparator | 4–5, 14–15 |
| Objectives | 10 | Specific objectives related to benefits and harms | 5, 10, 15 |
| **Methods: Patient and public involvement, trial design** | | | |
| Patient and public involvement | 11 | Details of, or plans for, patient or public involvement in the design, conduct, and reporting of the trial | 14 (participant input on app design) |
| Trial design | 12 | Description of trial design including type of trial, allocation ratio, and framework (e.g., superiority, equivalence, non-inferiority, exploratory) | 2, 5–6 |
| **Methods: Participants, interventions, and outcomes** | | | |
| Trial setting | 13 | Settings (e.g., community, hospital) and locations (e.g., countries, sites) where the trial will be conducted | 5–6 |
| Eligibility criteria | 14a | Eligibility criteria for participants | 5–6 |
| | 14b | If applicable, eligibility criteria for sites and for individuals who will deliver the interventions (e.g., surgeons, physiotherapists) | 7–8 (therapist role described; eligibility criteria not specified) |
| Intervention and comparator | 15a | Intervention and comparator with sufficient details to allow replication including how, when, and by whom they will be administered | 7–8 (online supplemental file 2, tables S8–S16) |
| | 15b | Criteria for discontinuing or modifying allocated intervention/comparator for a trial participant | 7–8, 13–14 (online supplemental file 2, tables S7–S8 and S15) |
| | 15c | Strategies to improve adherence to intervention/comparator protocols, and any procedures for monitoring adherence | 8 (online supplemental file 2, table S8) |
| | 15d | Concomitant care that is permitted or prohibited during the trial | 8 |
| Outcomes | 16 | Primary and secondary outcomes, including the specific measurement variable, analysis metric, method of aggregation, and time point for each outcome | 8–12 |
| Harms | 17 | How harms are defined and will be assessed (e.g., systematically, non-systematically) | 10, 13–14 (definitions and follow-up described; ascertainment method not specified) |
| Participant timeline | 18 | Time schedule of enrollment, interventions, assessments, and visits for participants. A schematic diagram is highly recommended | 5–8, 24 (Figure 1 and legend) |
| Sample size | 19 | How sample size was determined, including all assumptions supporting the sample size calculation | 10–11 |
| Recruitment | 20 | Strategies for achieving adequate participant enrollment to reach target sample size | 5–6 |
| **Methods: Assignment of interventions** | | | |
| Randomization: Sequence | 21a | Who will generate the random | 6 (REDCap method described; |

| | | | |
|---|---|---|---|
| generation | | allocation sequence and the method used | responsible person not named) |
| Randomization: Type | 21b | Type of randomization (simple or restricted) and details of any factors for stratification | 6 |
| Allocation concealment mechanism | 22 | Mechanism used to implement the random allocation sequence, describing any steps to conceal the sequence until interventions are assigned | 6 |
| Implementation | 23 | Whether the personnel who will enroll and those who will assign participants to the interventions will have access to the random allocation sequence | 6 (sequence concealed until allocation; access by role not specified) |
| Blinding | 24a | Who will be blinded after assignment to interventions (e.g., participants, care providers, outcome assessors, data analysts) | 6 |
| | 24b | If blinded, how blinding will be achieved and description of the similarity of interventions | 6, 8 (shared procedures described; blinding procedures not specified) |
| | 24c | If blinded, circumstances under which unblinding is permissible, and procedure for revealing a participant's allocated intervention during the trial | 6 (no unblinding planned) |
| **Methods: Data collection, management, and analysis** | | | |
| Data collection methods | 25a | Plans for assessment and collection of trial data, including processes to promote data quality and description of trial instruments with their reliability and validity | 8–10, 12 |
| | 25b | Plans to promote participant retention and complete follow-up, including outcome data collected for participants who discontinue or deviate from intervention protocols | 8, 13–14 (safety follow-up described; efficacy outcomes after withdrawal not specified) |
| Data management | 26 | Plans for data entry, coding, security, and storage, including processes to promote data quality | 12 |
| Statistical methods | 27a | Statistical methods used to compare groups for primary and secondary outcomes, including harms | 11–12 |
| | 27b | Definition of who will be included in each analysis (e.g., all randomized participants), and in which group | 11 |
| | 27c | How missing data will be handled in the analysis | 11–12 |
| | 27d | Methods for any additional analyses (e.g., subgroup and sensitivity analyses) | 11–12 |
| **Methods: Monitoring** | | | |
| Data monitoring committee | 28a | Composition of DMC; summary of its role; statement of independence; or explanation of why a DMC is not needed | 14 (site monitoring described; DMC or rationale not reported) |
| | 28b | Explanation of any interim analyses and stopping guidelines | 12, 14 (no interim analyses; trial stopping criteria) |
| Trial monitoring | 29 | Frequency and procedures for monitoring trial conduct. If there is | 14 (regular on-site visits; interval not specified) |

| | | | |
|---|---|---|---|
| | | no monitoring, give explanation | |
| **Ethics** | | | |
| Research ethics approval | 30 | Plans for seeking research ethics committee/institutional review board approval | 17 |
| Protocol amendments | 31 | Plans for communicating important protocol modifications to relevant parties | 17 (ethics committee described; other parties not specified) |
| Consent or assent | 32a | Who will obtain informed consent or assent from potential trial participants or authorized proxies, and how | 6, 17 (written consent; person obtaining it not specified) |
| | 32b | Additional consent provisions for collection and use of participant data and biological specimens in ancillary studies | 17 (exploratory blood sampling covered by main trial consent) |
| Confidentiality | 33 | How personal information about potential and enrolled participants will be collected, shared, and maintained to protect confidentiality | 6, 12, 18 |
| Ancillary and post-trial care | 34 | Provisions, if any, for ancillary and post-trial care, and for compensation to those who suffer harm from trial participation | 13–14 (AE care and safety follow-up; compensation not specified) |

*We strongly recommend reading this checklist in conjunction with the SPIRIT 2025 Explanation and Elaboration and the SPIRIT 2025 Expanded Checklist for important clarifications on all the items. See www.consort-spirit.org

Citation: Chan A-W, Boutron I, Hopewell S, Moher D, Schulz KF, et al. SPIRIT 2025 statement: updated guideline for protocols of randomised trials. BMJ 2025;389:e081477.

© 2025 Chan A-W et al. This is an Open Access article distributed under the terms of the Creative Commons Attribution License (https://creativecommons.org/licenses/by/4.0/).
