## Supplement for "Effects of an exercise prescription system (ACTIVE) on physical function in community-dwelling older adults: protocol for a single-centre, assessor-blinded, parallel-group randomised controlled trial"

**Online supplemental file 2**

### ACTIVE decision rules

This supplement describes how ACTIVE converts baseline assessment results into structured labels and uses them to generate, deliver and adapt individualised, home-based multicomponent exercise prescriptions. It covers the full cycle of functional assessment inputs, decision rules, prescription generation, home-based exercise, perceived difficulty ratings, and progression and regression. ACTIVE uses knowledge-based rules fixed before the trial. Label assignment and feedback-based adjustments follow these rules, with no online learning or changes to thresholds during the trial.

#### 1 Decision rules

ACTIVE links clinical assessment, exercise prescription and delivery through the digital platform in a traceable cycle. Each prescription can be traced to the original measurements, structured labels, selected modules, initial difficulty, video content and subsequent feedback.

##### 1.1 Rule priorities

The system applies rules in a fixed order to prevent conflicts between labels.

Safety restrictions take priority. If the pain score is >3 on the numerical rating scale (NRS) or an adverse event (AE) occurs, the system stops sending exercises for that day and displays a safety alert in the clinician interface until a clinician clears it manually.

Trigger rules operate independently. Each positive label activates only the corresponding exercise module, leaving other modules unchanged. The left and right sides are labelled separately, and exercises targeting the weaker side are assigned to that side only. Each module for which a rating is submitted is maintained, progressed or regressed independently.

##### 1.2 Decision unit

The exercise module is the smallest unit of decision-making in ACTIVE. Prescriptions may include lower-limb strength, whole-body strength, balance, coordination, flexibility and core training. Difficulty is adjusted independently for each module.

#### 2 Baseline assessment and structured labels

Descriptive labels are used to explain the prescription rules. Measurements are compared with the relevant thresholds by age, sex and side, and prescriptions are assigned according to the results.

##### 2.1 Balance

**Table S1. Balance assessment rules**

| Test | Age (years) | Threshold for men | Threshold for women | Below threshold | Threshold met |
|---|---|---|---|---|---|
| Eyes-open single-leg stance | 60–69 | ≥28.7 s | ≥25.1 s | bal_eo_below_l/r | bal_eo_met_l/r |
| Eyes-open single-leg stance | 70–79 | ≥18.3 s | ≥11.3 s | bal_eo_below_l/r | bal_eo_met_l/r |
| Eyes-open single-leg stance | 80–85 | ≥5.6 s | ≥7.4 s | bal_eo_below_l/r | bal_eo_met_l/r |
| Eyes-closed single-leg stance | 60–69 | ≥3.1 s | ≥2.5 s | bal_ec_below_l/r | bal_ec_met_l/r |

| Test | Age (years) | Threshold for men | Threshold for women | Below threshold | Threshold met |
|---|---|---|---|---|---|
| Eyes-closed single-leg stance | 70–79 | ≥1.9 s | ≥2.2 s | bal_ec_below_l/r | bal_ec_met_l/r |
| Eyes-closed single-leg stance | 80–85 | ≥1.3 s | ≥1.4 s | bal_ec_below_l/r | bal_ec_met_l/r |

Thresholds are based on the mean of three trials for each age and sex group in table 1 of Springer et al.[1] For participants aged 80–85 years, the reference values for adults aged 80–99 years in the original study are used.

#### 2.2 Coordination

Coordination is assessed using three functional movements. The labels dev0 to dev3 indicate that 0–3 types of movement quality deviations were observed. The deviations are trunk lean (trunk_lean), pelvic tilt (pelvic_tilt) and lower-limb alignment deviation (alignment_deviation). The left and right sides are assessed separately for the eyes-open single-leg stance and the single-leg sit-down. The labels dev2 and dev3 correspond to the same training level. The lowest level determines the final training difficulty. The coordination training levels, in ascending order of difficulty, are level_1 (beginner), level_2 (intermediate), level_3 (advanced) and level_4 (higher level).

**Table S2. Coordination assessment rules**

| Movement | Assessment result | Label | Coordination training level |
|---|---|---|---|
| Eyes-open single-leg stance | No deviations observed | sls_dev0_l/r | level_3 |
| Eyes-open single-leg stance | 1 type of deviation | sls_dev1_l/r | level_2 |
| Eyes-open single-leg stance | 2 types of deviation | sls_dev2_l/r | level_1 |
| Eyes-open single-leg stance | 3 types of deviation | sls_dev3_l/r | level_1 |
| Single-leg sit-down | Unable to complete | slsd_unable_l/r | level_1 |
| Single-leg sit-down | No deviations observed | slsd_dev0_l/r | level_4 |
| Single-leg sit-down | 1 type of deviation | slsd_dev1_l/r | level_3 |
| Single-leg sit-down | 2 types of deviation | slsd_dev2_l/r | level_2 |
| Single-leg sit-down | 3 types of deviation | slsd_dev3_l/r | level_2 |
| Squat jump | Unable to complete or NRS >3 | sj_limited | level_1 |
| Squat jump | No deviations observed | sj_dev0 | level_4 |
| Squat jump | 1 type of deviation | sj_dev1 | level_3 |
| Squat jump | 2 types of deviation | sj_dev2 | level_2 |
| Squat jump | 3 types of deviation | sj_dev3 | level_2 |

#### 2.3 Flexibility

**Table S3. Posterior chain flexibility**

| Age (years) | Reference threshold for men | Reference threshold for women |
|---|---|---|
| 60–64 | −6.35 cm | −1.27 cm |
| 65–69 | −7.62 cm | −1.27 cm |
| 70–74 | −8.89 cm | −2.54 cm |
| 75–79 | −10.16 cm | −3.81 cm |
| 80–84 | −13.97 cm | −5.08 cm |
| 85 | −13.97 cm | −6.35 cm |

Thresholds are the 25th percentiles from table 4 (women) and table 5 (men) of Rikli and Jones, converted using 1 inch = 2.54 cm.[2] For participants aged 85 years, the reference values for adults aged 85–89 years in the original study are used.

**Table S4. Lower-limb joint range of motion (ROM)**

| Movement | Assessment result | Label |
|---|---|---|
| Knee extension | Unable to reach full extension | knee_ext_limit_l/r |
| Knee flexion | Unable to reach full flexion, with pain on flexion | knee_flex_pain_l/r |
| Knee flexion | Unable to reach full flexion, with no pain on flexion | knee_flex_limit_l/r |
| Hip flexion | <120° | hip_flex_limit_l/r |
| Hip extension | Positive Thomas test | thomas_pos_l/r |

#### 2.4 Muscle strength

##### 2.4.1 Lower-limb muscle strength

Maximal isometric lower-limb strength is measured separately on each side. The strength asymmetry index (ASI) is calculated as the absolute difference in strength between sides divided by the strength of the stronger side and multiplied by 100%. Nasiri et al used the same formula in their prospective cohort study protocol.[3] Si et al used 15% to define groups in a study of isokinetic knee extensor strength.[4] Under the prespecified ACTIVE rules, an ASI >15% for a muscle group triggers targeted training of that muscle group on the weaker side.

Yang et al found that the isometric hamstrings-to-quadriceps strength ratio (H:Q) was associated with a history of falls in older adults. The optimal threshold for distinguishing participants with and without a fall history in their study was 0.733.[5] ACTIVE calculates the H:Q ratio separately for each side by dividing maximal isometric hamstring strength by maximal isometric quadriceps strength on the same side. The prespecified ACTIVE prescription rules assign hamstring training to the corresponding side when H:Q <0.80.

**Table S5. Lower-limb strength training rules**

| Muscle group | Trigger criteria | Label series |
|---|---|---|
| Quadriceps | ASI >15% and weaker side | quad_weak_l/r |
| Hamstrings | ASI >15% and weaker side | ham_weak_l/r |
| Gluteus medius | ASI >15% and weaker side | glutmed_weak_l/r |
| Hip adductors | ASI >15% and weaker side | add_weak_l/r |
| Gluteus maximus | ASI >15% and weaker side | glutmax_weak_l/r |

The table shows the ASI-based triggers. Hamstring training may also be triggered by an H:Q ratio <0.80 on the same side.

##### 2.4.2 Whole-body muscle strength

Thresholds for handgrip strength, appendicular skeletal muscle mass index (ASMI) measured using bioelectrical impedance analysis (BIA), and five-times sit-to-stand time were based on the 2025 Asian Working Group for Sarcopenia (AWGS) consensus.[6] ACTIVE applies the numerical gait speed threshold from the AWGS 2019 consensus to 4-m gait speed. The consensus uses a 6-m walk test. The threshold for the Short Physical Performance Battery (SPPB) total score was also based on the AWGS 2019 consensus.[7] Whole-body strength training is triggered when any of the five measures meets the criterion in table S6. This rule is used to assign exercise prescriptions.

**Table S6. Whole-body strength training rules**

| Measure | Trigger criteria | Label |
|---|---|---|
| Handgrip strength | Men aged 60–64 years, <34 kg, or aged ≥65 years, <28 kg<br>Women aged 60–64 years, <20 kg, or aged ≥65 years, <18 kg | grip_low |
| Appendicular skeletal muscle mass index | Men aged 60–64 years, <7.6 kg/m², or aged ≥65 years, <7.0 kg/m²<br>Women, <5.7 kg/m² | asmi_low |
| 4-m gait speed | <1.0 m/s | gait_low |
| Five-times sit-to-stand test | Aged 60–64 years, ≥10 s<br>Aged ≥65 years, ≥12 s | chair_slow |
| SPPB total score | ≤9 points | sppb_low |

#### 2.5 Cardiorespiratory endurance assessment and aerobic exercise education

Predicted 6-min walk distance (6MWD) is calculated for each participant using the equations of Enright and Sherrill.[8] ACTIVE compares the measured distance with the predicted distance. If the measured distance is below the predicted value, the system generates the cardio_edu label and delivers an aerobic exercise education video. Otherwise, the system assigns the cardio_edu_no label. Education videos are excluded from required training and the prescribed exercise dose.

Predicted 6MWD for men (m) = 7.57 × height (cm) − 5.02 × age (years) − 1.76 × body mass (kg) − 309.

Predicted 6MWD for women (m) = 2.11 × height (cm) − 2.29 × body mass (kg) − 5.78 × age (years) + 667.

### 3 Generation of the initial prescription

#### 3.1 Screening for risks and contraindications

**Table S7. Safety restrictions and alternative actions**

| Safety event or label | System restriction | Alternative action |
|---|---|---|
| NRS pain score >3 | Suspend exercise | Alert in the clinician interface. Exercise resumes after review. |
| AE during the intervention | Suspend exercise | Alert in the clinician interface. Exercise resumes after review. |

#### 3.2 Exercise dose and schedule

**Table S8. Exercise dose and schedule**

| | Prespecified rules |
|---|---|
| Intervention period | The intervention lasts 8 weeks. The final assessment is scheduled for the end of intervention week 8 (figure 1, study week W9 ± 3 days). For participants whose intervention is extended by 1 week under the protocol, the final assessment moves to study week W10. |

| | Prespecified rules |
|---|---|
| Training frequency | Strength training is performed every other day. Balance, coordination and flexibility training are performed daily. |
| Daily dose | The control group is prescribed a fixed daily dose of 7–12 exercises.<br>In the ACTIVE group, the prescribed daily exercise duration depends on baseline assessment results and may be adjusted in response to training feedback. |
| Difficulty feedback | ACTIVE participants rate perceived difficulty on a five-point scale after completing the relevant exercise module (table S15). |
| Reminders and monitoring | Training reminders are sent daily at 06:00.<br>The system records daily video completion rates for each module and check-in frequency. |
| Adherence support | If the daily completion rate is below 60%, the clinician interface generates an alert and the study team follows up by telephone. |
| Intervention extension | During the 8-week intervention, if the average completion rate is below 30% in any week, the intervention is extended by 1 week and the final assessment is postponed accordingly. |

### 4 Exercise library and prescribed doses

#### 4.1 Balance training

**Table S9. Balance exercises**

| Difficulty | Exercise | Dose |
|---|---|---|
| Eyes-open balance, beginner | Supported eyes-open single-leg stance | 20 s × 3 sets |
| Eyes-open balance, advanced | Unsupported eyes-open single-leg stance | 20 s × 3 sets |
| Eyes-closed balance, beginner | Supported eyes-closed single-leg stance | 10 s × 3 sets |
| Eyes-closed balance, advanced | Unsupported eyes-closed single-leg stance | 10 s × 3 sets |

#### 4.2 Lower-limb strength training

**Table S10. Lower-limb strength exercises**

| Difficulty | Exercise | Dose |
|---|---|---|
| Quadriceps, beginner | Seated knee extension with a yoga block | 10–15 repetitions × 3 sets |
| Quadriceps, intermediate | Seated resisted knee extension | 10–15 repetitions × 3 sets |
| Quadriceps, advanced | Wall sit | 3–4 sets |
| Quadriceps, higher level | Single-leg sit-to-stand | 6–10 repetitions × 1 set |
| Hamstrings, beginner | Resisted knee flexion in a forward-leaning position | 10–15 repetitions × 3 sets |
| Hamstrings, intermediate | Standing resisted hip extension | 10–15 repetitions × 3 sets |
| Hamstrings, advanced | Single-leg glute bridge | 10–15 repetitions × 3 sets |
| Gluteus medius, beginner | Clamshell | 10–15 repetitions × 3 sets |
| Gluteus medius, intermediate | Side-lying leg raise | 10–15 repetitions × 3 sets |
| Gluteus medius, advanced | Lateral step-up | 10–15 repetitions × 3 sets |
| Hip adductors, beginner | Seated heel-to-knee touch | 10–15 repetitions × 3 sets |
| Hip adductors, intermediate | Standing hip adduction | 10–15 repetitions × 3 sets |

| Difficulty | Exercise | Dose |
|---|---|---|
| Hip adductors, advanced | Standing hip adduction with a resistance band | 10–15 repetitions × 3 sets |
| Gluteus maximus, beginner | Prone hip extension with the knee flexed | 10–15 repetitions × 3 sets |
| Gluteus maximus, intermediate | Single-leg glute bridge | 10–15 repetitions × 3 sets |
| Gluteus maximus, advanced | Single-leg sit-to-stand | 6–10 repetitions × 1 set |

#### 4.3 Whole-body strength training

**Table S11. Whole-body strength exercises**

| Difficulty | Exercise | Dose |
|---|---|---|
| Beginner | Wall sit | 3–4 sets |
| | Abdominal curl-up | 10–15 repetitions × 1 set |
| | Biceps curl | 10–15 repetitions × 3 sets |
| Intermediate | Two-leg sit-to-stand | 10–15 repetitions × 3 sets |
| | Standing push-up | 10–15 repetitions × 3 sets |
| | Military press | 10–15 repetitions × 3 sets |
| Advanced | Single-leg sit-to-stand (both sides) | 6–10 repetitions × 1 set |
| | Kneeling push-up | 10–15 repetitions × 3 sets |
| | Single-leg glute bridge (both sides) | 10–15 repetitions × 3 sets |

#### 4.4 Coordination training

**Table S12. Coordination exercises**

| Difficulty | Exercise | Dose |
|---|---|---|
| Beginner | Two-leg squat | 10–15 repetitions × 3 sets |
| | Supported eyes-open single-leg stance | 20 s × 3 sets |
| Intermediate | Unsupported eyes-open single-leg stance | 20 s × 3 sets |
| | Wall-supported lunge | 10–15 repetitions × 3 sets |
| Advanced | Single-leg squat | 6–10 repetitions × 3 sets |
| | Squat with lateral pressure against a yoga block | 6–10 repetitions × 3 sets |
| Higher level | Landing drill | 6–10 repetitions × 3 sets |
| | Jump | 6–10 repetitions × 3 sets |

#### 4.5 Flexibility training

**Table S13. Flexibility exercises**

| Category | Exercise | Dose |
|---|---|---|
| Posterior chain flexibility | Seated toe reach | 20–30 s × 2 sets |
| Knee extension ROM | Seated toe reach | 20–30 s × 2 sets |
| | Calf stretch | 20–30 s × 2 sets |

| Category | Exercise | Dose |
|---|---|---|
| Knee flexion ROM (painful) | Dynamic knee mobilisation | 20–30 s × 2 sets |
| Knee flexion ROM (pain-free) | Quadriceps foam rolling | 40–60 s × 2 sets |
| | Side-lying anterior thigh stretch | 20–30 s × 2 sets |
| Hip flexion ROM | Lying 90/90 (beginner) | 12–20 repetitions × 1 set |
| | Hand-supported 90/90 (intermediate) | 12–20 repetitions × 1 set |
| | Unsupported 90/90 (advanced) | 12–20 repetitions × 1 set |
| Hip extension ROM | Iliopsoas stretch | 20–30 s × 2 sets |
| | Quadriceps foam rolling | 40–60 s × 2 sets |
| | Side-lying anterior thigh stretch | 20–30 s × 2 sets |

##### 4.6 Core training

**Table S14. Core exercises**

| Difficulty | Exercise | Dose |
|---|---|---|
| Beginner | Seated abdominal draw-in | 10–15 repetitions × 3 sets |
| | Static dead bug | 10–15 repetitions per side |
| Intermediate | Kneeling plank | 10–15 repetitions |
| | Kneeling side plank | 10–15 repetitions per side |
| Advanced | Dynamic dead bug | 10–15 repetitions × 3 sets |
| | Standard side plank | 10–15 repetitions |

#### 5 Progression and regression based on training feedback

Participants rate their perceived difficulty on a five-point scale after completing each exercise module. The system uses the current and previous ratings for the same module to update its difficulty for the next training day according to prespecified rules.

**Table S15. Rules for the five-point perceived difficulty scale**

| Rating | Perceived difficulty | Trigger | Decision for the next training day |
|---|---|---|---|
| 1 | Very easy | One rating | Increase the module difficulty by one level |
| 2 | Easy | Two consecutive ratings | Increase the module difficulty by one level |
| 3 | Moderate | One rating | Maintain the current level |
| 4 | Difficult | Two consecutive ratings | Decrease the module difficulty by one level |
| 5 | Very difficult | One rating | Decrease the module difficulty by one level |

When the highest difficulty level in the exercise library is reached, the system maintains that level. At the lowest difficulty level, a rating of ‘very difficult’ automatically triggers an alert in the clinician interface. Progression and regression are applied independently to each exercise module. Changes to one module leave the other modules unchanged. Safety restrictions, movement quality and pain take priority over progression.

#### 6 Standardised multicomponent training for the control group

The control group completes 8 weeks of home-based exercise through the same WeChat mini programme, following a fixed set of exercises and a standardised schedule. Strength training is performed every other day, and balance, coordination and flexibility training are performed daily. Baseline assessments do not determine exercise content, and individual feedback does not alter exercise difficulty.

**Table S16. Fixed exercises, schedule and prescribed doses for the control group**

| Training module | Fixed exercise | Schedule | Dose |
|---|---|---|---|
| Balance | Supported eyes-open single-leg stance (left and right) | Every training day | 20 s × 3 sets |
| Coordination | Two-leg squat | Every training day | 10–15 repetitions × 3 sets |
| Hip mobility | Hand-supported 90/90 | Every training day | 12–20 repetitions × 1 set |
| Knee mobility | Side-lying anterior thigh stretch (left and right) | Every training day | 20–30 s × 2 sets |
| Lower-limb strength | Side-lying leg raise (left and right) | Strength training days | 10–15 repetitions × 3 sets |
| Lower-limb strength | Seated heel-to-knee touch (left and right) | Strength training days | 10–15 repetitions × 3 sets |
| Whole-body strength | Standing push-up | Strength training days | 10–15 repetitions × 3 sets |
| Core strength | Static dead bug | Strength training days | 10–15 repetitions × 3 sets |

Health education includes videos on aerobic exercise for cardiorespiratory health and educational content on diet and nutrition.

### References


1. Springer BA, Marin R, Cyhan T, et al. Normative values for the unipedal stance test with eyes open and closed. J Geriatr Phys Ther 2007;30:8–15. doi:10.1519/00139143-200704000-00003
2. Rikli RE, Jones CJ. Functional fitness normative scores for community-residing older adults, ages 60–94. J Aging Phys Act 1999;7:162–81. doi:10.1123/japa.7.2.162
3. Nasiri Z, Letafatkar A, Hosseinzadeh M. Investigating the association between intralimb strength ratio, interlimb strength and range of motion asymmetry index, and functional limitations with fall incidence among older adults: protocol for a prospective cohort study. BMJ Open Sport Exerc Med 2024;10:e002110. doi:10.1136/bmjsem-2024-002110
4. Si B, Zhu H, Wei X, et al. The mechanism of static postural control in the impact of lower limb muscle strength asymmetry on gait performance in the elderly. PeerJ 2024;12:e17626. doi:10.7717/peerj.17626
5. Yang F, Simpkins C, Trammell AR, et al. Hamstrings-quadriceps strength ratio could be related to falls in community-living older adults. J Electromyogr Kinesiol 2024;77:102900. doi:10.1016/j.jelekin.2024.102900
6. Chen LK, Hsiao FY, Akishita M, et al. A focus shift from sarcopenia to muscle health in the Asian Working Group for Sarcopenia 2025 Consensus Update. Nat Aging 2025;5:2164–75. doi:10.1038/s43587-025-01004-y
7. Chen LK, Woo J, Assantachai P, et al. Asian Working Group for Sarcopenia: 2019 consensus update on sarcopenia diagnosis and treatment. J Am Med Dir Assoc 2020;21:300–7.e2. doi:10.1016/j.jamda.2019.12.012
8. Enright PL, Sherrill DL. Reference equations for the six-minute walk in healthy adults. Am J Respir Crit Care Med 1998;158:1384–7. doi:10.1164/ajrccm.158.5.9710086
